# Exploring the associations between screen time and sleep quality in adult residents of Ahmedabad: A cross-sectional study

**DOI:** 10.1101/2024.11.11.24315299

**Authors:** Bharat Patel, Disha Pandya, Bhadrawati Prajapati, Omkumar Doshi

## Abstract

**Background and rationale:** Sleep is essential for overall health, but many people experience disruptions in their sleep patterns due to busy schedules and technology use. This study aims to examine the impact of screen time on sleep quality, considering factors such as smoking, caffeine use, napping, physical activity, and stress levels.

**Methodology:** We enrolled 371 participants from Ahmedabad for a cross-sectional study. 278 participants were included after the screening. They completed a questionnaire covering demographics, screen time, covariates, stress using the PSS10 scale, and sleep quality using the PSQI scale. Data was analyzed using IBM SPSS Statistics 22 after entering it into MS Excel.

**Result:** This study examines the relationships between lifestyle factors and sleep quality among adults aged 18 to 40 in Ahmedabad. The findings suggest that increased screen time (r = 0.518, p < 0.001) and higher stress levels (r = 0.530, p < 0.001) are associated with lower sleep quality. Additionally, nap frequency and physical activity show significant associations with sleep ratings. However, no significant connections were found between caffeine use at night or smoking/tobacco usage and sleep quality.

**Conclusion:** The Spearman correlation indicates a moderate correlation between screen time and sleep quality, as well as between stress and sleep quality. The chi-square test shows significant associations between screen time before bed, napping, and physical activity with sleep quality. However, there is no association between nighttime caffeine consumption or smoking/tobacco use and sleep quality.

## 1. Introduction

Sleep is a fundamental physiological activity that is critical to general health and well-being. Sleep consistency and quality have been identified as critical variables in defining an individual’s physical, mental, and emotional well-being. However, in today’s world of hectic work schedules, social obligations, and the pervasive influence of technology, many people face interruptions in their sleep patterns, resulting in irregular sleep cycles[1]. Sleep is divided into two neurophysiological phases recorded by electroencephalogram (EEG) and electromyogram (EMG): rapid eye movement (REM) and non-rapid eye movement (NREM)[2]. Humans experience between 2 and 8 such cycles every night, with the most usual pattern being 4-5 cycles of 90-minute periods[3]. The EEG essentially shifts when a person goes to sleep from a condition of high frequency and low voltage waves indicative of a waking state to a state of higher voltage and slower waves indicative of NREM sleep[4]. NREM sleep is classified into three stages: Stage 1 is the relaxed wakefulness stage, which lasts less than 10 minutes and is characterized by slow breathing, heartbeats, and relaxation of the muscles. Stage 2 represents light sleep, with a slower heart rate and breathing, as well as increased muscular relaxation and slower brain activity. This stage lasts around 30-60 minutes. Stage 3 also called slow wave sleep (SWS) or deep sleep, during which breathing and heartbeats become extremely slow. Muscles are relaxed at this stage, and brain waves are even slower. This stage lasts between 20 and 40 minutes. Following NREM sleep, there is the REM phase, which is indicated by increased frequency and lower voltage action, in the REM phase, breathing and heartbeats are rapid and most dreaming occurs[4, 5]. During NREM sleep, sympathetic tone decreases and parasympathetic activity increases, resulting in a state of diminished activity. Increased parasympathetic activity and regulated sympathetic activity characterize REM sleep[4].

Irregular sleep cycles include a variety of sleep problems, such as irregular bedtimes, disrupted sleep-wake rhythms, and changes in sleep length. These interruptions are frequently caused by lifestyle choices, occupational pressures, or underlying health issues[6]. Several studies have found that irregular sleep patterns have negative consequences on various elements of health. According to research, irregular sleep patterns are associated with an increased risk of acquiring chronic disorders such as obesity, diabetes, cardiovascular disease, and reduced immunological function[7]. Circadian rhythm disruption, hormone imbalance changes, and neurotransmitter system disruptions may all contribute to the negative health effects seen in those with irregular sleep habits[8, 9]. While much research has been done on the effects of irregular sleep cycles, there is still a need for more research into the particular processes by which irregular sleep impacts numerous aspects of health. This thesis seeks to add to the existing body of knowledge by investigating the complex relationship between irregular sleep cycles and their numerous implications on physical, mental, and emotional well-being[10].

Digital gadgets and online scenarios are seen as essential components of the contemporary generation’s lives. Self-luminous gadgets that emit blue and bright light in the evening and at night may interfere with the circadian rhythm by inhibiting the generation of melatonin or changing when it is generated and retained and this can lead to decreased drowsiness, difficulty initiating sleep, and non-restorative sleep. The blue region of the spectrum centered around 450–480 nm, is where the circadian photoreceptor system reaches its maximum sensitivity. This explains why blue light is so effective in suppressing melatonin and boosting alertness. The majority of contemporary computers, televisions, tablets, smartphones, and a growing number of lightbulbs for homes are powered by LEDs with a peak wavelength in the ∼460 nm blue region. The pineal gland generates melatonin, also known as N-acetyl-5-methoxy tryptamine, an endogenous hormone that is exclusively produced at night and plays a role in circadian rhythm and the human sleep-wake cycle. Supplementing with exogenous melatonin is well tolerated and does not seem to have any negative short- or long-term consequences. It has been demonstrated that melatonin synchronizes circadian cycles and enhances the onset, duration, and quality of sleep. It is crucial for enhanced neuronal functioning, sleep modulation, circadian rhythm maintenance, and free-radical scavenging. Melatonin, then, provides a less harmful alternative to the prescription treatments for sleep problems that are now on the market. Excess screen time leads to sedentary behaviour and increased mortality and morbidity[11–14].

Caffeine use reduces subsequent total sleep time, sleep onset delay, waking after sleep onset, sleep efficiency, and sleep architecture. Specifically, the closer to bedtime higher levels of caffeine are ingested, the greater the reduction in overall sleep duration[15]. Physical activity holds an important position in affecting sleep quality, with plenty of studies revealing the dual-way link between exercise and sleep-wake circadian oscillations. Research shows that incidences of difficulty in sleeping or insomnia at the current point in time can indicate that a person is more likely to develop more sedentary behaviour in the future. It is now known that exercise is a factor a better sleep quality, but there is no sufficient evidence whether the opposite is true or not: better sleep has the effect of increased physical activity level[16]. Persons, who practice exercise regularly, tend to sleep better than those, who do it rarely or even never altogether. When it comes to acute physical activity it could only play a small role in sleep quality and duration, at the same time if you moderate exercise regularly it can be very helpful in extending sleep duration and improving sleep quality, and also it could speed up the process of falling asleep i.e. sleep onset[17]. For a properly appointed workout, it is essential to consider the timing of it. It’s noticeable that the aerobic workouts performed in the morning provide perks to sleeping quality that are greater than similar actions in the afternoon or evening[18]. Nicotine, the main component of cigarettes, affects sleep by creating disruptions before bedtime and midnight desires. It also causes abnormal circadian cycles, snoring, and obstructive sleep apnea (OSA). Sleep problems, including trouble falling asleep, are frequent among smokers of all ages[19]. These sleep disturbances are caused by nicotine’s influence on the sleep-wake cycle neuron[20]. Stress can disrupt sleep because activation of the hypothalamus pituitary adrenal (HPA) and/or sympathetic nerve systems leads to wakefulness, and these substances include corticotropin-releasing hormone (CRH), adrenocorticotropic hormone (ACTH), cortisol or corticosterone, noradrenaline, and adrenaline. Chronic stress causes a rise in distal corticosteroids and disrupts sleep[21].

**Figure 1:**
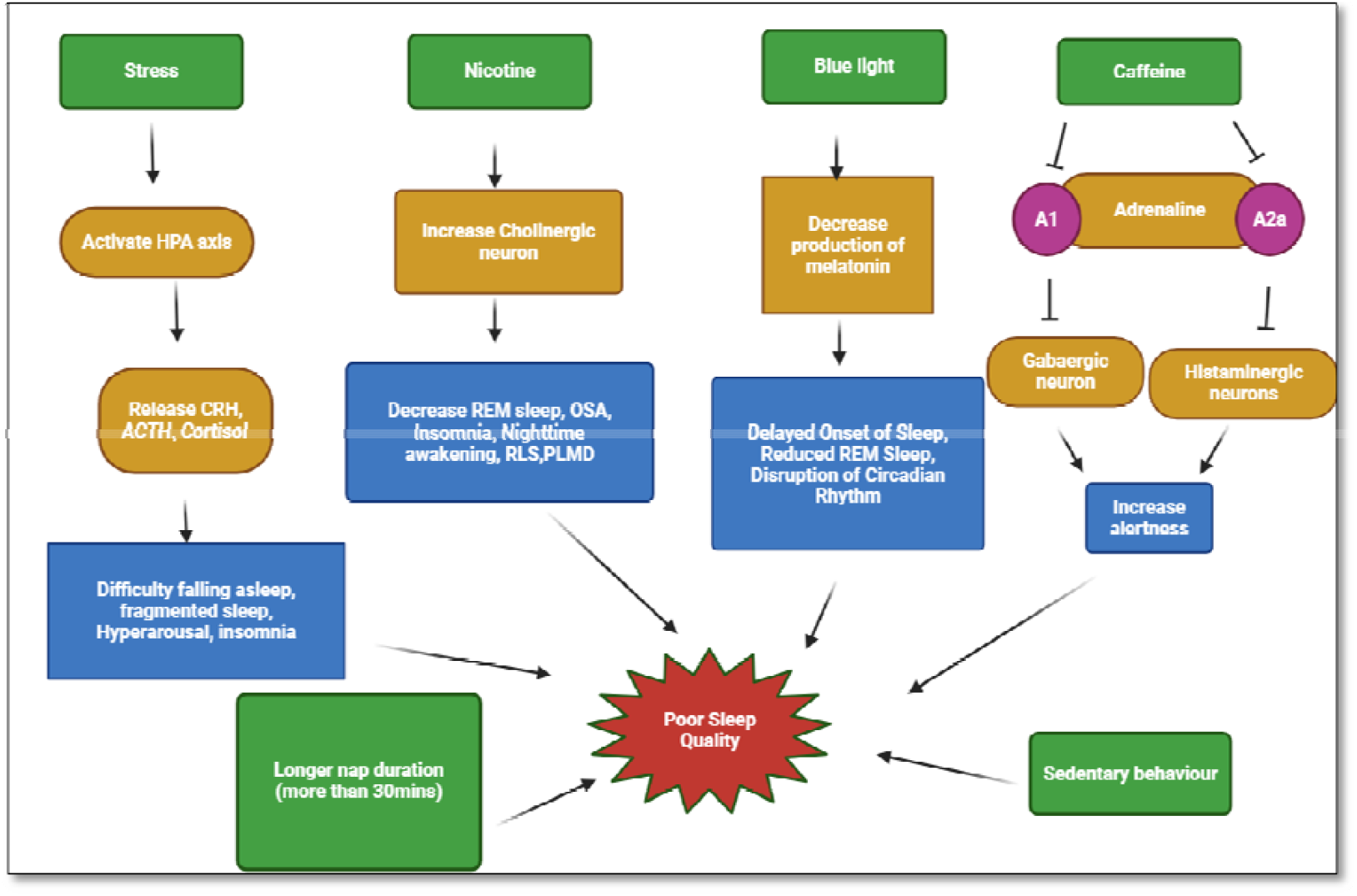
Factors affecting sleep quality. The following factors have been found to impact the quality of sleep significantly: 1) Stress, which stimulates the secretion of cortisol, has been shown to have a negative effect on sleep quality. 2) Nicotine, which increases cholinergic neurons, has also been linked to poor sleep quality. 3) Blue light has been found to decrease melatonin production and delay the onset of sleep. 4) Caffeine, which blocks adrenaline receptors A1 and A2a, can increase alertness and negatively impact sleep quality. In addition, longer nap durations (more than 30 minutes) and sedentary behavior have also been associated with poor sleep quality.

## 2. Aim and Objective

- To study the impact of screen time, physical activity, and lifestyle factors such as caffeine consumption, and tobacco consumption on sleep quality in Ahmedabad adults.

### Objectives

- To determine the association between screen time and sleep quality.
- To investigate the relationship between stress and sleep quality.
- To determine the association between other factors like physical activity, caffeine consumption, tobacco products, and, sleep quality.

## 3. Methodology

This cross-sectional observational study was conducted among adult residents of Ahmedabad, aged 18 to 40 years, to explore the associations between screen time and sleep quality. The study, titled “Exploring the Associations between Screen Time and Sleep Quality in Adult Residents of Ahmedabad: A Cross-Sectional Study,” received ethical approval from the institutional ethics committee (LMIEC/2023-24/06). Participants were recruited based on specific inclusion criteria, including residency in Ahmedabad, the ability to comprehend the study’s purpose, willingness to provide informed consent, and regular use of electronic devices (smartphones, laptops, tablets, computers, or television) for at least 15 minutes daily. Exclusion criteria included the use of sleep-related medications, treatment for chronic diseases, night-shift workers, individuals with irregular work schedules impacting sleep, and participants with a recent history of substance abuse. A minimum sample size of 256 was required.

Data collection occurred from September 2023 to March 2024, utilizing both online (via Google Forms) and offline (physical forms) methods. The Google Form was distributed through email and social media, and a QR code was also made available. Participants were first provided with an informed consent form (ICF) in their preferred language, followed by a screening form. Based on the screening form, individuals were either included or excluded from the study. Participants who met the inclusion criteria completed a demographic questionnaire and provided self-reported responses on variables such as screen usage, caffeine consumption, naps, physical activity, and smoking habits. The questionnaire included multiple-choice, closed-ended questions. Sleep quality was assessed using the Pittsburgh Sleep Quality Index (PSQI), while stress levels were evaluated using the Perceived Stress Scale (PSS-10).

Data were entered into MS Excel, and statistical analysis was performed using IBM SPSS Statistics 22. The Kolmogorov-Smirnov test was employed to determine the distribution of data, whether parametric or non-parametric. Spearman’s rank correlation was used to examine the associations between screen time, sleep quality, and stress. Chi-square tests were performed to evaluate the significant associations between sleep quality and various variables, including naps, physical activity, screen time before bed, and nighttime caffeine use. Fisher’s exact test was applied to examine the relationship between sleep quality and smoking or tobacco use.

## 4. Results

**Table 1:** Demonstrates the distribution of study participants by background characteristic.

| Sociodemographic profile | N (%) |
| --- | --- |
| Gender |  |
| Male | 138(49.64) |
| Female | 140(50.36) |
| Zone of Ahmedabad |  |
| East Zone | 29(10.43) |
| West Zone | 137(49.28) |
| North Zone | 22(7.91) |
| South Zone | 27(9.71) |
| North West | 28(10.07) |
| South West | 29(10.43) |
| Central Zone | 06(2.15) |
| Screen time per day(hr) |  |
| 1 | 11(3.97) |
| 2 | 29(10.43) |
| 3 | 31(11.15) |
| 4 | 36(12.95) |
| 5 | 40(14.39) |
| 6 | 46(16.55) |
| 7 | 29(10.43) |
| 8 | 20(7.19) |
| 9 | 12(4.31) |
| 10 and more than 10 | 24(8.63) |
| Screen time in bed before sleep(min) |  |
| Less than 30min | 118(42.45) |
| More than 30 min | 160(57.55) |
| Taking Nap |  |
| Yes | 135(48.56) |
| No | 143(51.44) |
| Smoking |  |
| Yes | 11(3.96) |
| No | 256(92.09) |
| Prefer not to answer | 11(3.96) |
| Night time caffeine |  |
| Yes | 58(20.86) |
| No | 220(79.14) |
| Physical activity |  |
| Yes | 121(43.53) |
| No | 157(56.47) |
| Stress |  |
| Low (0-13) | 48(17.27) |
| Moderate (14-26) | 213(76.62) |
| High (27-40) | 17(6.12) |
| Sleep score |  |
| Good (0-5) | 177(63.66) |
| Poor (6-21) | 101(36.33) |

In the study, 278 participants were enrolled among which 138(49.64%) were male and 140(50.36%) were females. Out of 278 participants, 29 were from the east zone, 137 were from the west zone, 22 were from the north zone, 27 were from the south zone, 28 were from the northwest zone, 29 were from the southwest zone, and 6 were from the central zone of Ahmedabad. The use of screen time per day was estimated through the questionnaire of each participant. Maximum screen time usage was found to be 4,5,6 hours per day respectively 12.95%, 14.39%, and 16.55%. Out of 278 participants, 57.55% were using screen time more than 30 minutes (screen time in bed before sleep) and 42.45% were using less than 30 mins. Out of 278 participants, 48.56% were taking naps in the daytime and 57.55% were not taking naps. Of 278 participants, 3.96% were current smokers, 3.96% were not answering the questions and 92.09% were non-smokers. Out of 278 participants, 20.86% consume caffeine at night and 79.14% do not consume caffeine at night. Out of 278 participants, 43.53% were performing physical activity and 56.47% were not performing physical activity. Out of 278 participants 17.27% had low stress levels, 76.62 had moderate and 6.12% had high levels of stress. Out of 278 participants, 36.33% of participants had poor sleep scores and 63.66% of participants had good sleep scores.

**Table 2:** Kolmogorov-Smirnov test (at confidence interval (CI) 95%, α= 0.05)

|  | Sleep score | Stress | Screen time | ST before bed | Smoking | Caffeine | Nap | Physical Activity |
| --- | --- | --- | --- | --- | --- | --- | --- | --- |
| Median | 5.0 | 18.0 | 5.0 | 1.0 | 0.0 | 0.0 | 0.0 | 0.0 |
| IQR | 3 | 6 | 4 | 1 | 0 | 0 | 1 | 1 |
| Sig. | <0.001 | <0.001 | <0.001 | <0.001 | <0.001 | <0.001 | <0.001 | <0.001 |
|  | Sleep score | Stress | Screen time | ST before bed | Smoking | Caffeine | Nap | Physical Activity |
| Median | 5.0 | 18.0 | 5.0 | 1.0 | 0.0 | 0.0 | 0.0 | 0.0 |
| IQR | 3 | 6 | 4 | 1 | 0 | 0 | 1 | 1 |
| Sig. | <0.001 | <0.001 | <0.001 | <0.001 | <0.001 | <0.001 | <0.001 | <0.001 |
|  | Sleep score | Stress | Screen time | ST before bed | Smoking | Caffeine | Nap | Physical Activity |
| Median | 5.0 | 18.0 | 5.0 | 1.0 | 0.0 | 0.0 | 0.0 | 0.0 |
| IQR | 3 | 6 | 4 | 1 | 0 | 0 | 1 | 1 |
| Sig. | <0.001 | <0.001 | <0.001 | <0.001 | <0.001 | <0.001 | <0.001 | <0.001 |

Our data was found to be non-parametric as per the Kolmogorov-Smirnov test. Screen time before bed was divided in two: Less than 30 minutes which was coded by 0 and more than 30 minutes coded by 1. Likewise in nap no nap was coded as 0 and taking a nap was coded as 1. In smoking, not smoking was coded as 0, smoking as 1, and prefer not to answer as 2. In nighttime caffeine, if yes then coded as 1 and no then 0. In physical activity, if yes then code was1 and no then 0. In sleep score, if the score was ≤5 coded as 0, and more than 5 coded as 1.

**Table 3:** Spearman’s rank correlation.

| Variable | Correlation coefficient |
| --- | --- |
| Screen time and sleep | 0.518 |
| Stress and sleep | 0.530 |

Spearman’s rank correlation coefficient test was performed to check the correlation between sleep score and screen time and also for sleep score and stress. The Spearman rank correlation test found a moderate association between screen time and sleep (r = 0.518, p < 0.001). This conclusion implies that there is a significant relationship between screen time and sleep.

The Spearman rank correlation test revealed a moderate association between the stress and sleep score (r = 0.530, p < 0.001), indicating a statistically significant relationship between the stress and sleep score.

**Table 4:** Association of global Pittsburgh sleep quality index score with various factors (n=278)

| Variables | Group | PSQI score | | | $\chi^2_{(0.05,1)}$ | p-value |
| --- | --- | --- | --- | --- | --- | --- |
|  |  | Poor, n (%) | Good, n (%) | Total, n (%) |  |  |
| Gender | Male | 46 (33.33) | 92 (66.67) | 138 (49.64) | 1.064 | 0.302 |
|  | Female | 55 (39.25) | 85 (60.71) | 140 (50.36) |  |  |
| Screen time before bed | Less than 30 min | 29 (24.58) | 89 (75.42) | 118 (42.45) | 12.247 | 0.000 |
|  | More than 30 min | 72 (45) | 88 (55) | 160 (57.55) |  |  |
| Taking nap | Yes | 59 (43.70) | 76 (56.30) | 135 (48.56) | 6.167 | 0.013 |
|  | No | 42 (29.37) | 101 (70.63) | 143 (51.44) |  |  |
| Physical activity | Yes | 35 (28.93) | 86 (71.07) | 121 (43.53) | 5.079 | 0.024 |
|  | No | 66 (42.04) | 91 (57.96) | 157 (56.47) |  |  |
| Caffeine consumption at night | Yes | 25 (43.10) | 33 (56.90) | 58 (20.86) | 1.453 | 0.228 |
|  | No | 76 (34.55) | 144 (65.45) | 220 (79.14) |  |  |

According to the findings, out of a sample of 138 males, 33.33% exhibit poor sleep quality. Similarly, the study reveals that among the 140 females surveyed, 39.25% reported poor sleep quality. As shown from the chi-square result (χ^2^ _(0.05,1)_ = 12.247, p < 0.001), there was a significant association between the variables “screen time before bed” and “sleep quality.” Based on a chi-square test result of 6.167 and a significance threshold of 0.013, the null hypothesis of no effect of “nap” on “sleep quality” may be rejected. This shows that there is a statistically significant relationship between the two variables. The p-value of 0.013 is less than the specified significance threshold of 0.05, indicating that the observed discrepancy between observed and predicted cell counts is unlikely to be due to chance. The chi-square test found a significant association between the variables “physical activity” and “sleep quality” (χ^2^ _(0.05,1)_ = 5.079, p = 0.024). This finding suggests that there is a link between physical activity and sleep ratings among research participants. Based on a chi-square test result of 1.453 and a significance level of 0.228, the null hypothesis of no effect of “use of caffeine at night” on “sleep quality” cannot be rejected. This means that there is no statistically significant relationship between the two variables. The p-value of 0.228 is larger than the significance level of 0.05, indicating that the observed difference between observed and predicted cell counts is unlikely to be due to chance. As a result, the findings do not support the study question or hypothesis that coffee intake does not influence sleep scores.

**Table 5:**
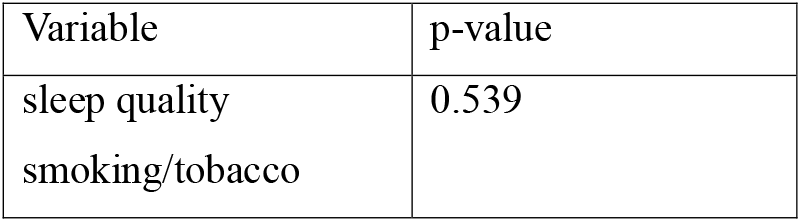
Fisher’s exact test for sleep quality and use of smoking/tobacco.

According to Fisher’s exact test, the P value was found 0.539, the null hypothesis that “smoking/tobacco” does not influence “sleep quality” cannot be rejected. This indicates that there is no statistically significant association between the two variables. As a result, the data do not support the study’s hypothesis that smoking/tobacco usage does not affect sleep quality.

## 5. Discussion

The study investigates the complex relationship between lifestyle factors and sleep quality in Ahmedabad’s adult population. Notably, characteristics such as longer screen exposure, increased stress, nap frequency, physical activity levels, and screen time before bedtime all have statistically significant relationships with sleep quality. However, the study found no significant relationships between nocturnal coffee consumption or smoking/tobacco use and sleep ratings. These findings emphasize the importance of comprehensive therapies focused on altering lifestyle behaviours to improve overall sleep health.

Several studies have reported similar findings regarding the detrimental effects of excessive screen time on sleep patterns. Research by Hale and Guan found that prolonged screen time, particularly engagement with electronic devices such as smartphones and computers, was associated with reduced sleep duration and increased sleep disturbances among children and adolescents [22]. Additionally, a Study by Cain & Gradisar highlighted the negative impact of screen exposure before bedtime on sleep onset latency (SOL) and overall sleep quality, consistent with the observed association between screen time and sleep scores in our study[23]. Similarly, the significant correlation between stress levels and sleep quality observed in our study is consistent with previous research indicating that higher stress levels are associated with poorer sleep outcomes[24–26]. Stress and sleep are two interconnected aspects of our lives. High levels of stress can negatively impact the quality and duration of sleep, while poor sleep can further exacerbate stress levels. This reaffirms the notion that psychological well-being plays a critical role in regulating sleep patterns and it is important to find ways to manage stress and establish healthy sleep habits to maintain overall well-being. There are significant associations between physical activity and sleep quality and nap and sleep quality.

On the other hand, the lack of significant associations between nocturnal caffeine consumption or smoking/tobacco usage and sleep quality diverges from some previous studies. While research by Drake et al. and Clark & Landolt has reported negative effects of caffeine intake on sleep quality, our findings suggest limited direct effects of caffeine or smoking/tobacco use on sleep quality within the studied demographic. This may indicate variations in individual susceptibility or cultural differences in caffeine and tobacco consumption patterns and quantities, warranting further investigation[27, 28]. Previous research suggests that higher rate of poor sleep quality in smokers, but smoking is not associated with worsened sleep quality or daytime sleepiness[29]. While Dugas and colleagues discovered that heavier smokers with higher levels of nicotine dependency were more likely to have poor sleep quality, Himal et al. observed no statistically significant link between smoking behaviour (e.g., cigarettes/day) and sleep quality[30].

## 6. Conclusion

Overall, the findings of this study indicate a robust relationship between screen usage and sleep quality among Ahmedabad residents aged 18 to 40. Individuals who spent more time using screens, such as smartphones, tablets, and laptops, had lower sleep quality. This is a significant result, given the ubiquitous use of screens in today’s society and the possible harmful influence on people’s sleep quality. Our study found a moderate link between screen time and sleep quality, suggesting that excessive screen exposure may have a negative impact on sleep patterns. Similarly, stress levels have a similar relationship, showing that higher stress levels are associated with lower sleep quality. Furthermore, substantial connections are shown between nap frequencies, physical activity, and sleep ratings, highlighting the relevance of daytime activities in determining nocturnal sleep. Notably, screen time before bed is identified as a key factor influencing sleep quality, emphasizing the importance of attentive screen usage behaviours before bedtime. However, the study found no significant link between caffeine use at night or smoking/tobacco use and sleep quality. While these findings may indicate that these variables may not have a direct impact on sleep quality in this community, more study is needed to investigate any indirect effects or variances among populations. In conclusion, this study emphasizes the multidimensional nature of factors impacting sleep quality in adult Ahmedabad residents, as well as the necessity of supporting good lifestyle practices to promote overall sleep health. More research on the intricate relationship between lifestyle variables and sleep quality is needed to create tailored therapies for better sleep outcomes.

## 7. Limitations of this study

### 1. Self-Report Measures

The use of self-reported data, particularly for screen time, coffee intake, physical activity, and smoking/tobacco use, may result in recollection bias or social desirability bias. Participants may underreport or overreport their behaviours, resulting in mistakes in the data.

### 2. Study Design

The longitudinal form of the study can provide the capacity to demonstrate causal correlations between variables. The study can include examining how screen time, stress levels, or sleep quality changed over time.

### 3. Confounding factors

Factors such as socioeconomic background, work status, and medical history may impact the relationship between screen time, stress, and sleep quality might have affected the outcome but were not adequately addressed.

## Data Availability

All data produced in the present work are contained in the manuscript

## Acknowledgment

The authors are grateful to Prof. GBS, Department of Pharmacology, L. M. College of Pharmacy, Ahmedabad, Gujarat, India for support and guidance.

## Author contribution

All authors contributed equally.

